# Development and validation of neurological health score using machine learning algorithms

**DOI:** 10.64898/2026.02.11.26346101

**Authors:** Sandhya Kiran Pemmasani, Shravya Atmakuri, R G Shakthiraju, Anuradha Acharya

**Affiliations:** Mapmygenome India Limited

**Keywords:** CRISPR, Wheat, RUBY, Copy number, *Agrobacterium*, T-DNA

## Abstract

Neurological health score (NHS), indicating the health of brain and nervous system, helps in identifying high risk individuals, and in recommending lifestyle modifications. In the present study, we developed NHS based on genetic, lifestyle and biochemical variables associated with eight neurological disorders - dementia, stroke, Parkinson’s disease, amyotrophic lateral sclerosis, schizophrenia, bipolar disorder, multiple sclerosis and migraine. UK Biobank data from Caucasian individuals was used to develop the model, and the data from individuals of Indian ethnicity was used to validate the model. Logistic regression and XGBoost algorithms were used in selecting the significant variables for the disorders. NHS developed from the selected variables was found to be very significant after adjusting for age and sex (AUC:0.6, OR: 0.95). Higher NHS was associated with a lower risk of neurological disorders and better social well-being. Highest NHS group (top 25%) showed 1.3 times lower risk compared to the rest of the individuals. Results of our study help in developing a framework for quantifying the neurological health in clinical setting.

## INTRODUCTION

Neurological health refers to the functioning of the brain and nervous system, which can be identified by cognitive ability, emotional stability, social well-being and neurological disorders like dementia, stroke, Parkinson’s disease, amyotrophic lateral sclerosis, schizophrenia, migraine etc. It contributes significantly to the overall health and quality of life. Genetic, lifestyle (exercise, diet, sleep, social connections) and biochemical factors (cholesterol, triglycerides, glucose) influence neurological health. Genome-wide association studies (GWAS) have identified several genetic variants associated with neurological disorders. But the disorders are multifactorial, caused by interactions between genetics and environmental factors. For example, in a recent study, Jiang et al [1] observed that healthy lifestyle can mitigate the genetic susceptibility to Parkinson’s disease. Similar observations were made in the case of Alzheimer’s and dementia [2,3]. Singh et al [4] developed a 21-point Brain Care Score that can predict the risk of dementia and stroke. They focused on factors that can realistically be modified by the patient or practitioner. Other risk factors such as age, genetics, education level, and socio-economic status were not considered. Integration of genetic, environmental and lifestyle factors helps in understanding the onset, in personalizing the treatments and in taking preventive measures.

Machine learning (ML) algorithms, such as XGBoost, random forests, logistic regression are being increasingly used in clinical applications to accurately predict the disease risk. Especially, in the context of integrating clinical and genetic data, ML models show superior performance by handling non-linear relationships among the variables. Yan et al [5] used XGBoost and logistic regression to predict the postoperative 5-year survival in patients with glioma, and found that XGBoost has better performance in predicting the risk of death compared to multivariate logistic regression. Rahmatinejad et al [6] worked on various ML algorithms, and concluded that XGBoost achieved highest precision, sensitivity and accuracy in predicting in-hospital mortality rates. Gu et al [7] observed that machine learning-based sarcopenia prediction model provides a valuable decision support tool for clinical screening and intervention of sarcopenia. But the major limitation of XGBoost is the lack of interpretability. It does not provide statistical significance of the variables. It can be combined with logistic regression to understand the effect sizes.

In the present study, we developed a neural health score that quantifies the health of brain and nervous system using data from UK Biobank, a prospective study of 500,000 participants aged 40-69 years, recruited between 2006 and 2010 [8]. We used logistic regression to construct an interpretable risk model, and trained a gradient-boosted decision tree model (XGBoost) as a complementary analysis to assess whether non-linear interactions improved discrimination. We validated the model on Indian participants of UK Biobank. To our knowledge, this is the first study on neural health that integrates genetic, lifestyle and biochemical factors. NHS can be used in clinical setting to identify high risk individuals, and to recommend lifestyle modifications.

## METHODOLOGY

### Study Data

UK Biobank is a large population-based prospective cohort study of 500,000 participants, aged 40-69 years when recruited in 2006-2010, living in the United Kingdom [11]. Extensive phenotypic and genotypic data of the participants was collected across four assessment visits. In this study, we used genetic, lifestyle, sociodemographic and biochemical variables associated with eight neurological disorders - dementia, stroke, Parkinson’s disease, amyotrophic lateral sclerosis (ALS), multiple sclerosis, schizophrenia, bipolar disorder and migraine. Case/control status was ascertained based on International Classification of Diseases (ICD) code 10 (Field ID#: 41270), ICD9 (Field ID#: 41271) and self-report (Field ID#: 20002). Data fields and codes used for the neurological disorders are given in Table 1. Age at diagnosis was taken as earliest of doctor diagnosed age (Field ID#: 2976), self-reported age (Field ID#: 20009), first in-patient diagnosis in ICD10 records (Field ID#: 41280) and ICD9 records (Field ID#: 41281). Participants were excluded if the age at diagnosis was ess than the age at baseline assessment or if the information was not available on age at diagnosis.

**Table 1.** Codes used for disease ascertainment.

| Disease | ICD10 | ICD9 | Self Report |
| --- | --- | --- | --- |
| Dementia | F00, F01, F02, F03, G30 | 2901 | 1263 |
| Stroke | I60, I61, I63, I64 | 430, 431, 434, 436 | 1081, 1086, 1491, 1583 |
| Amyotrophic Lateral Sclerosis (ALS) | G12.2 | 3352 | 1259 |
| Bipolar disorder | F31 | 296 [2960, 2961, 2962, 2963, 2964, 2965, 2966, 2969] | 1291 |
| Schizophrenia | F20 | 295 [2950, 2951, 2953, 2954, 2955, 2956, 2957, 2959] | 1289 |
| Parkinson's Disease | G20 | 3320 | 1262 |
| Multiple sclerosis | G35 | 3409 | 1261 |
| Migraine | G43 | 346 [3460, 3461, 3462, 3468, 3469] | 1265 |

UK Biobank has information on 30 blood related biochemical parameters like ALD, ASP, albumin, vitamin D etc. Variables that have data on at least one lakh participants were only included in the study. Lifestyle variables included physical activity, current smoking status, alcohol intake frequency, diet, BMI, sleep duration, stress, friends and family visits, walking pace etc. Socio-demographic variables included age, gender, education, employment status, income, Townsend deprivation index and hand grip strength. Data fields and codes are given in Table 2.

**Table 2.** Codes of the variables used in the study.

| Variable | UK Biobank Field ID | Variable Description |
| --- | --- | --- |
| Ethnicity | 21000 | 1001: Caucasian; 3001: Indian |
| BMI | 21001 | Body mass index (BMI) – Kg/m<sup>2</sup> |
| Diet |  | Total score of 4 categories (Salt, fruits & Veg, Meat and whole grains) was considered. |
| Alcohol Intake Frequency | 1558 | Alcohol Intake Frequency |
| Smoking Status | 20116 | Smoking status. |
| Sleep Duration | 1160 | Sleep duration |
| Sleeplessness | 1200 | Sleeplessness / insomnia |
| Stress | 2070 | Frequency of tenseness / restlessness in last 2 weeks. |
| Friends Family Visits | 1031 | Frequency of friend/family visits. Categorized into Almost daily, 2-3 times a week etc. |
| Hearing loss | 2247 | Hearing loss |
| Loneliness | 2020 | Often feel lonely ? |
| Confide | 2110 | Able to confide in someone close |
| Physical Activity |  | Number of minutes of physical activity per week |
| Walking Pace | 924 | Slow, Average, Brisk walking |
| Paid Employment | 6142 | Employment status. Categorized into paid employed, retired, unemployed, part time etc. |
| Income | 738 | Average total household income before tax. |
| Hand Grip Strength | Left hand: 46<br>Right hand: 47 | Kg (Average of left and right hands was taken) |
| LDL (mg/dL) | 30780 | LDL Direct (mmol/L) multiplied by 38.67 |
| HDL (mg/dL) | 30760 | HDL (mmol/L) multiplied by 38.67 |
| Total Cholesterol (mg/dL) | 30690 | Total cholesterol (mmol/L) multiplied by 38.67 |
| Triglycerides (mg/dL) | 30870 | Triglycerides (mmol/L) multiplied by 88.57 |
| Systolic Blood Pressure |  |  |
| HbA1C (%) | 30750 | Glycated haemoglobin (mmol/mol)<br>$HbA1c (\%) = (HbA1c (mmol/mol) / 10.93) + 2.15$ |
| IGF1 (nmol/L) | 30770 | Insulin-like Growth Factor 1 |
| ApoA (mg/dL) | 30630 | Apolipoprotein A (g/L) multiplied by 100 |
| ApoB | 30640 | Apolipoprotein B |
| LipoproteinA (mg/dL) | 30790 | Lipoprotein A (nmol/L) |
| CRP (mg/L) | 30710 | C-reactive protein (mg/L) |
| Creatinine (mg/dL) | 30700 | Creatinine (umol/L) divided by 88.4 |
| Urate (mg/dL) | 30880 | Urate (umol/L) divided by 59.48 |
| Urea (mg/dL) | 30670 | Blood Urea (mmol/L) multiplied by 6.01 |
| Cystatin C | 30720 | Cystatin C (mg/L) |
| Total Protein (g/dL) | 30860 | Total Protein (g/L) divided by 10 |
| Vitamin D (ng/mL) | 30890 | Vitamin D (nmol/L) divided by 2.5 |
| Total Bilirubin (mg/dL) | 30840 | Total Bilirubin (umol/L) divided by 17.1 |
| Albumin (g/L) | 30600 | Albumin (g/L) |
| AST (U/L) | 30650 | Aspartate Aminotransferase |
| ALP (U/L) | 30610 | Alkaline phosphatase |
| ALT (U/L) | 30620 | Alanine aminotransferase |
| GGT (U/L) | 30730 | Gamma glutamyltransferase (U/L) |
| Calcium (mg/dL) | 30680 | Calcium (mmol/L) multiplied by 4.008 |
| Phosphate (mg/dL) | 30810 | Phosphate (mmol/L) multiplied by 3.1 |

For each of the neurological disorder, GWAS catalog was screened to identify genetic variants associated with the disorder. Minimal curation was done to filter out variants based on availability of odds ratio, presence on GSA-V3 chip and GWAS p-value threshold of 5x10^-8^. When there are multiple studies for an SNP, the study with largest sample size was taken. Presence on GSA-V3 was considered for enabling the portability of health score in clinical settings. Polygenic risk scores (PRS) were calculated as weighted sum of odds ratios using PLINK2.0 software. Imputed genotype data (Field ID#: 22828) available in BGEN v1.2 format was used in the analysis. QCTOOL v2 [17] was used to retrieve the samples and variants of interest. Variants with INFO score >=0.3 were considered in calculating the PRS. Number of SNPs considered for each disorder is given in Table 3

**Table 3.** Numbers of SNPs used in developing polygenic risk score.

| Neurological Disorder | Number of SNPs |
| --- | --- |
| Dementia | 11 |
| Stroke | 10 |
| Amyotrophic Lateral Sclerosis (ALS) | 8 |
| Parkinson's disease | 66 |
| Bipolar disorder | 30 |
| Schizophrenia | 183 |
| Multiple Sclerosis | 193 |
| Migraine | 36 |

### Statistical analyses

Correlations were calculated among the variables, and only one variable was retained among the correlated variables. Pearson’s correlation coefficient threshold of 0.9 was used for continuous variables, and Cramer’s V threshold of 0.3 was used for categorical variables. Logistic regression model was used to identify variables significantly associated with each of the disorders. XGBoost classifier was used as an add-on analysis to check non-linear effects and interactions among the variables. Variables that were significant in any of the disorders were finally considered for building an overall model for neurological disorders. Individuals with any of the eight neurological disorders were marked as cases while building the overall model.

Values of the each variable considered in the final model were divided into distinct categories, and each category was given a score. Neurological health score (NHS) was calculated as sum of scores of all the variables. Here, the assumption was that individuals with higher score carry lower risk of developing any disorder. Logistic regression model was built on NHS along with age and sex. Caucasian data was divided into 80-20 split for training and testing. Indian data was used to validate the model. Accuracy of the model was assessed using area under curve (AUC) from receiver operating curves (ROC). Analyses were done with PLINK2.0 and R v4.2.

## RESULTS

Supplementary Table 1 gives the characteristics of UK Biobank cohort for the variables considered in the study. A total of 147,781 participants with complete data on all the variables were included in the final analysis. Correlation analysis indicated that LDL cholesterol was correlated with total cholesterol and ApoA with coefficient values greater than 0.9. Similarly, HDL cholesterol was correlated with total cholesterol and ApoB, and total cholesterol was correlated with ApoB. So, we removed HDL, LDL and ApoA, and retained total cholesterol and ApoA. Income variable was removed due to high correlation with Employment status.

In the case of dementia, a total of 147,724 participants were considered in the analysis after excluding 57 prevalent cases. 1,340 incident dementia cases (0.9%) were recorded in the follow-up period. Logistic regression model and SHAP plots are given in Supplementary Information. Logistic regression analysis indicated that stress, income status, hand grip strength, HbA1C, AST and ALT levels are significantly associated with dementia at p-value <0.001. As expected, PRS for dementia is highly significant. SHAP plot indicated that PRS has highest significance after the Age variable. Though loneliness increases the risk for dementia, hearing loss doesn’t play any role in increasing the dementia risk. Table 4 shows the summary of the variables that are significant for various disorders. Walking pace, Stress and loneliness are significant in five of eight disorders. Similarly, employment status is significant in four disorders.

**Table 4.** Variables significant for various disorders.

| Variable | Dementia | Stroke | ALS | BPD | SCZ | PD | MS | MGN |
| --- | --- | --- | --- | --- | --- | --- | --- | --- |
| Age | √ | √ | √ |  |  | √ | √ |  |
| Male Gender | √ | √ | √ |  |  | √ |  | √ |
| BMI |  |  |  | √ |  |  |  | √ |
| Diet_Score1 |  |  |  |  |  |  |  |  |
| Diet_Score2 |  |  |  |  |  | √ |  |  |
| Alcohol Intake Frequency | √ |  |  |  | √ |  |  | √ |
| Current Smoker |  | √ |  | √ |  |  | √ |  |
| Sleep Duration |  |  |  |  |  |  |  |  |
| Sleeplessness | √ | √ |  |  |  | √ |  | √ |
| Stress | √ |  |  | √ | √ | √ |  | √ |
| Friends Family Visits |  | √ |  | √ |  |  |  |  |
| Physical Activity |  |  |  |  |  |  |  |  |
| Walking Pace1 | √ | √ |  |  |  |  | √ | √ |
| Walking Pace2 | √ | √ |  |  |  | √ | √ | √ |
| Paid Employment | √ |  |  | √ | √ | √ |  |  |
| Hearing loss |  |  |  | √ |  |  |  | √ |
| Loneliness | √ | √ |  | √ |  |  | √ | √ |
| Confide | √ |  |  |  |  |  |  |  |
| HandGripStrength | √ |  |  |  |  | √ |  |  |
| total cholesterol | √ | √ |  |  |  |  |  |  |
| Triglycerides | √ |  |  | √ |  | √ |  |  |
| Systolic blood pressure |  | √ |  | √ |  |  |  | √ |
| HbA1C | √ | √ |  |  |  |  |  |  |
| IGF1 | √ |  | √ |  |  | √ |  |  |
| ApoA |  | √ |  |  |  |  |  |  |
| CRP |  |  |  |  |  | √ |  |  |
| Creatinine | √ | √ |  |  |  |  |  | √ |
| Urate |  |  |  |  |  |  |  |  |
| Urea |  |  |  |  |  |  |  |  |
| CystatinC | √ | √ |  |  |  |  |  |  |
| Total Protein |  |  |  |  |  |  |  |  |
| VitaminD | √ | √ |  |  |  |  | √ |  |
| Total Bilirubin |  |  |  |  |  |  |  |  |
| Albumin | √ | √ |  |  | √ |  |  |  |
| AST | √ | √ |  |  |  |  |  |  |
| ALP |  |  |  |  |  |  |  |  |
| ALT | √ | √ |  |  |  | √ |  |  |
| GGT |  | √ |  |  |  | √ |  |  |
| Calcium | √ | √ |  |  |  |  |  |  |
| Phosphate |  |  |  | √ |  |  |  |  |
| PRS_Dementia | √ |  |  |  |  |  |  |  |
| PRS_Stroke |  |  |  |  |  |  |  |  |
| PRS_ALS |  |  |  |  |  |  |  |  |
| PRS_BPD |  |  |  |  |  |  |  |  |
| PRS_SCZ |  |  |  |  |  |  |  |  |
| PRS_PD |  |  |  |  |  |  |  |  |
| PRS_MS |  |  |  |  |  |  | √ |  |
| PRS_MGN |  |  |  |  |  |  |  |  |

### Combination of Neurological disorders

Incident cases of any of the eight neurological disorders were considered as cases for calculating risk for overall neurological health. The cumulative incidence rate is 9%, with 13,303 cases out of 147,781 participants. Table 5 gives the results of logistic regression model built on the training data. As expected, PRS of the disorders came out as the most significant variables. Among the lifestyle variables, stress, income and loneliness are the most significant variables. Among the biochemical variables, cystatinC, vitamin D, ALT and GGT are the most significant variables. Figure 1 gives SHAP plot on the same data. Alcohol intake frequency, employment, handgrip strength, walking pace and stress are the top five significant variables that increase the risk of neurological disorders. Figure 2 shows the variable importance.

**Table 5.** Logistic regression model built on training datas.

|  | Estimate | Std. Error | z value | Pr(>\|z\|) |
| --- | --- | --- | --- | --- |
| (Intercept) | -7.35E-01 | 3.09E-01 | -2.382 | 0.01720 * |
| Age | 2.40E-02 | 1.72E-03 | 13.991 | < 2e-16 *** |
| Male_Gender | -1.08E-01 | 3.41E-02 | -3.158 | 0.00159 ** |
| BMI | -1.00E-02 | 2.56E-03 | -3.923 | 8.73e-05 *** |
| Diet_Score1 | -3.67E-02 | 2.15E-02 | -1.709 | 0.08741 . |
| Diet_Score2 | -3.86E-02 | 2.57E-02 | -1.501 | 0.13347 |
| Alcohol_Intake_Frequency | -2.43E-01 | 2.04E-02 | -11.917 | < 2e-16 *** |
| currentSmoker | 3.25E-02 | 3.47E-02 | 0.937 | 0.34868 |
| SleepDuration | 2.34E-02 | 9.13E-03 | 2.56 | 0.01046 * |
| Sleeplessness | -3.08E-02 | 2.26E-02 | -1.363 | 0.17298 |
| Stress | 2.06E-01 | 2.21E-02 | 9.323 | < 2e-16 *** |
| Friends_Family_Visits | 1.53E-03 | 2.33E-02 | 0.066 | 0.94772 |
| Physical_Activity | -2.55E-02 | 1.97E-02 | -1.294 | 0.19565 |
| WalkingPace1 | -6.16E-01 | 3.60E-02 | -17.135 | < 2e-16 *** |
| WalkingPace2 | -7.07E-01 | 3.86E-02 | -18.307 | < 2e-16 *** |
| Paid_Employment | -2.60E-01 | 2.34E-02 | -11.13 | < 2e-16 *** |
| Hearing_loss | 1.05E-01 | 2.13E-02 | 4.924 | 8.48e-07 *** |
| Loneliness | -1.40E-01 | 2.60E-02 | -5.394 | 6.88e-08 *** |
| Confide | -1.82E-02 | 2.24E-02 | -0.814 | 0.41582 |
| HandGripStrength_Avg | -8.47E-03 | 1.36E-03 | -6.21 | 5.31e-10 *** |
| totChol | -3.02E-03 | 2.81E-04 | -10.75 | < 2e-16 *** |
| Triglycerides | 9.31E-06 | 1.42E-04 | 0.065 | 0.94784 |
| sysBP | -4.04E-05 | 5.76E-04 | -0.07 | 0.94399 |
| HbA1C | -1.40E-02 | 1.78E-02 | -0.786 | 0.43188 |
| IGF1 | 9.12E-04 | 1.79E-03 | 0.509 | 0.61068 |
| ApoA | -1.83E-03 | 4.67E-04 | -3.913 | 9.12e-05 *** |
| CRP | -5.58E-03 | 2.51E-03 | -2.228 | 0.02585 * |
| Creatinine | 2.36E-02 | 7.59E-02 | 0.31 | 0.7562 |
| Urate | -2.46E-02 | 9.76E-03 | -2.521 | 0.01169 * |
| Urea | -2.45E-03 | 1.36E-03 | -1.8 | 0.07192 . |
| CystatinC | 4.64E-01 | 8.50E-02 | 5.453 | 4.95e-08 *** |
| Total_Protein | -6.28E-02 | 3.00E-02 | -2.092 | 0.03641 * |
| VitaminD | -9.38E-03 | 1.19E-03 | -7.86 | 3.83e-15 *** |
| Total_Bilirubin | -4.87E-02 | 3.82E-02 | -1.276 | 0.20185 |
| Albumin | -1.14E-02 | 4.81E-03 | -2.361 | 0.01824 * |
| AST | 6.33E-03 | 1.44E-03 | 4.385 | 1.16e-05 *** |
| ALP | -1.05E-03 | 4.19E-04 | -2.494 | 0.01264 * |
| ALT | -6.45E-03 | 1.19E-03 | -5.421 | 5.94e-08 *** |
| GGT | 1.74E-03 | 2.58E-04 | 6.747 | 1.51e-11 *** |
| Calcium | 4.76E-02 | 3.21E-02 | 1.483 | 0.13809 |
| Phosphate | -1.20E-02 | 2.03E-02 | -0.594 | 0.55283 |
| PRS_Dementia | 6.59E-02 | 9.25E-03 | 7.127 | 1.03e-12 *** |
| PRS_Stroke | -1.56E-02 | 9.46E-03 | -1.651 | 0.09867 . |
| PRS_ALS | -8.00E-03 | 9.35E-03 | -0.855 | 0.39232 |
| PRS_BPD | -5.18E-03 | 9.40E-03 | -0.551 | 0.58152 |
| PRS_SCZ | 5.19E-03 | 9.42E-03 | 0.552 | 0.58123 |
| PRS_PD | -1.13E-02 | 9.31E-03 | -1.218 | 0.22321 |
| PRS_MS | 2.77E-02 | 9.34E-03 | 2.965 | 0.00302 ** |
| PRS_MGN | 8.62E-02 | 9.37E-03 | 9.202 | < 2e-16 *** |

**Figure 1.**
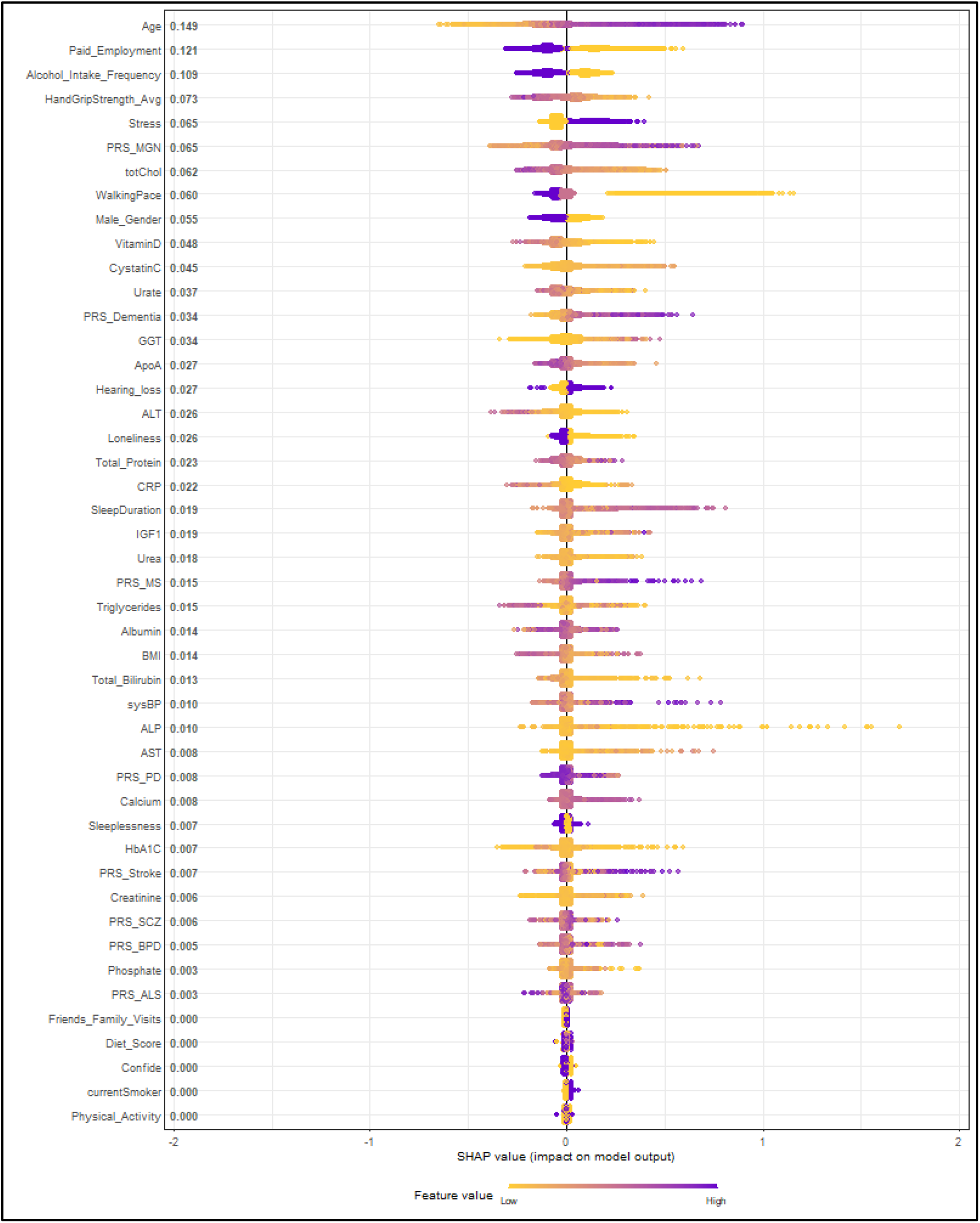
SHAP plot for combined Neurology

**Figure 2.**
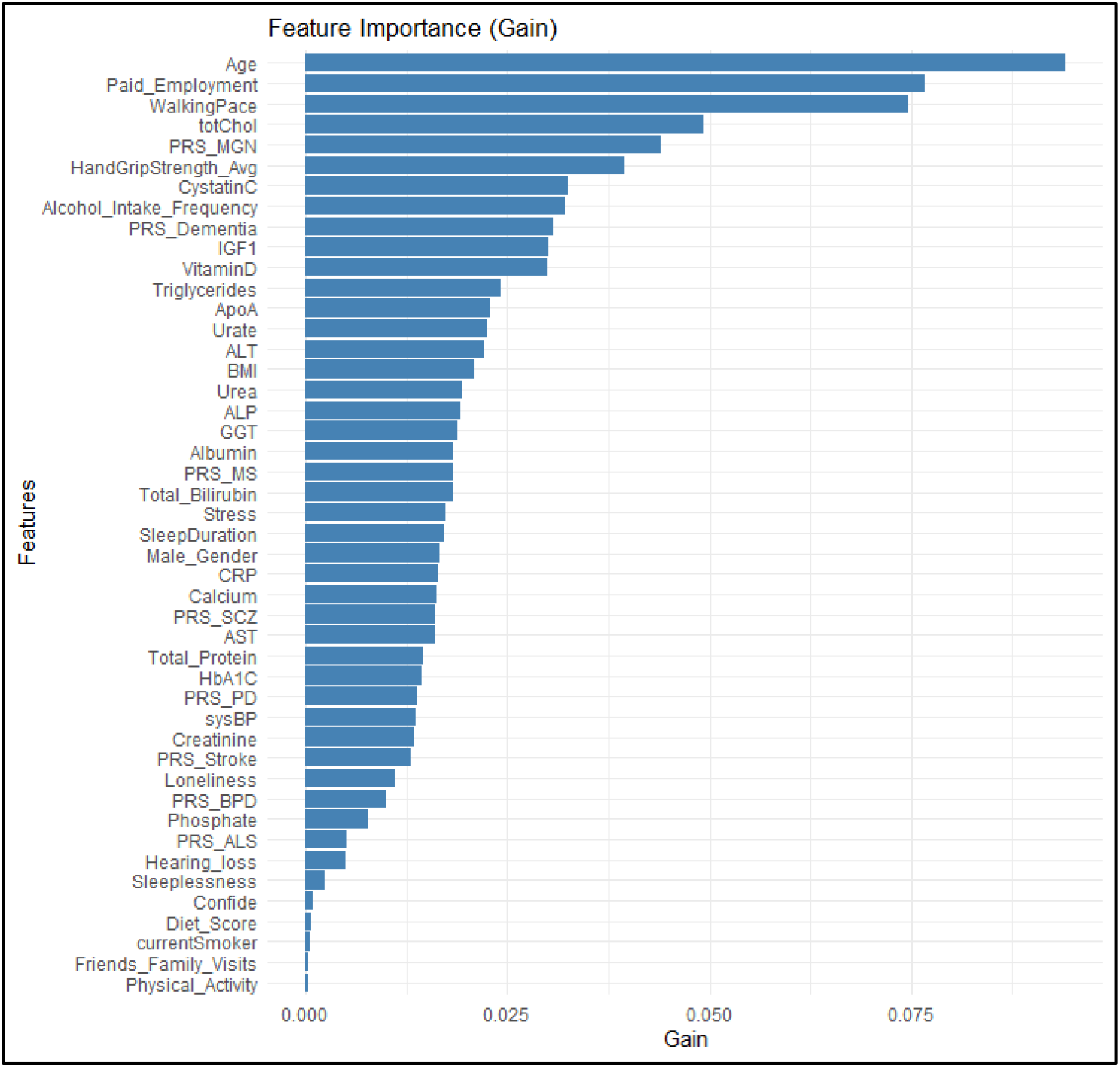
Feature importance

### Neurological Health Score (NHS)

Values of the each variable considered in the study were divided into distinct categories, and each category was given a score. Table 6 gives the score given for each category of the variables. Higher score was given for the category that decreases the risk of any disorder. Sum of the scores of all variables was considered as Neurological Health Score (NHS). Logistic regression model was built on NHS along with age and sex. Table 7 gives the odds ratios obtained for NHS when age and gender were added as covariates.

| Dataset | OR/SD | AUC |  |
| --- | --- | --- | --- |
|  |  | Age+Sex | NHS+Age+Sex |
| UK Biobank Caucasian | 0.95 | 0.58 | 0.6 |

## Data Availability

All data produced in the present study were obtained from UK Biobank

https://www.ukbiobank.ac.uk/

## Notes

### Competing Interest Statement

The authors have declared no competing interest.

### Funding Statement

This study did not receive any funding

### Author Declarations

Data was obtained from UK Biobank

